# Disease severity dictates SARS-CoV-2-specific neutralizing antibody responses in COVID-19

**DOI:** 10.1101/2020.07.29.20164285

**Authors:** Xiangyu Chen, Zhiwei Pan, Shuai Yue, Fei Yu, Junsong Zhang, Yang Yang, Ren Li, Bingfeng Liu, Xiaofan Yang, Leiqiong Gao, Zhirong Li, Yao Lin, Qizhao Huang, Lifan Xu, Jianfang Tang, Li Hu, Jing Zhao, Pinghuang Liu, Guozhong Zhang, Yaokai Chen, Kai Deng, Lilin Ye

## Abstract

COVID-19 patients exhibit differential disease severity after SARS-CoV-2 infection. It is currently unknown as to the correlation between the magnitude of neutralizing antibody (NAb) responses and the disease severity in COVID-19 patients. In a cohort of 59 recovered patients with disease severity including severe, moderate, mild and asymptomatic, we observed the positive correlation between serum neutralizing capacity and disease severity, in particular, the highest NAb capacity in sera from the patients with severe disease, while a lack of ability of asymptomatic patients to mount competent NAbs. Furthermore, the compositions of NAb subtypes were also different between recovered patients with severe symptoms and with mild-to-moderate symptoms. These results reveal the tremendous heterogeneity of SARS-CoV-2-specific NAb responses and their correlations to disease severity, highlighting the needs of future vaccination in COVID-19 patients recovered from asymptomatic or mild illness.

## Introduction

As of July 28, 2020, the pandemic of coronavirus disease 2019 (COVID-19), caused by severe acute respiratory syndrome coronavirus 2 (SARS-CoV-2) infection, has claimed 16,341,920 clinically confirmed cases and 650,805 deaths worldwide^1^. The infected patients show heterogeneous clinical manifestations, which can be generally classified into four groups, including severe, moderate, mild and asymptomatic, according to the severity of symptoms. Despite daily increasing confirmed cases and death, currently no medical agents are approved to prevent SARS-CoV-2 infection or treat COVID-19 patients.

A growing body of evidence shows that recovered COVID-19 patients can generate IgG-type antibodies specifically binding to various structure proteins of SARS-CoV-2 particles shortly after the onset of disease, albeit at variable levels^3-6^. Among these virus specific antibodies, only those capable of blocking SARS-CoV-2 spike (S) protein mediated viral attachment and/or entry of host cells, called neutralizing antibodies (NAbs), can effectively curtail infection^7^. The convalescent plasma or sera containing NAbs harvested from recovered patients have shown promising results in treating COVID-19 patients of critical illness in several small-scale clinic trials^8-11^. In addition, a variety of human monoclonal antibodies (mAbs) of potent SARS-CoV-2 neutralizing activities has been cloned from memory B cells from recovered COVID-19 patients^12-21^, holding great potentials for prophylactic or therapeutic use. However, little is known regarding the relationship between disease severity and the magnitude of SARS-CoV-2 -specific NAbs responses in patients recovered from COVID-19. Defining the association of disease severity to NAb responses will facilitate the screening of COVID- 19 recovered patients as therapeutic plasma donors as well as memory B cell providers for cloning high-affinity human neutralizing mAbs to prevent or treat COVID-19.

The circulation of high-titer NAbs provides the immediate protection against corresponding viral infections, which can be achieved by recovering from natural infection or by inducing from vaccine immunization. Thus far, there is no vaccine approved for COVID-19 prophylaxis, albeit several types of COVID-19 vaccines, including inactivated, vector-based, DNA and mRNA vaccines^-5^, are undergoing early stages of clinical trials. Additionally, the NAb titers can predict the possibility of re-infection in patients recovered from a primary viral infection. Currently, there are few clues regarding whether the patients recovered from COVID-19 can be protected from re-infection or will still require vaccination in the future when effective vaccines become available.

## Results

### Antibody responses to SARS-CoV-2 in COVID-19 recovered patients with different symptom severity

To explore the potential association between SARS-CoV-2 S protein-specific antibody responses and the disease severity in recovered COVID-19 patients, we included a cohort of 59 adult patients, 48 of mild (n=4), moderate (n=34) and severe (n=10) symptoms admitted to Guangzhou Eighth People’s Hospital and 11 asymptomatic adult patients admitted to Chongqing Public Health Center Hospital. The median age of these patients was 47 (33-62, interquartile range (IQR)) years old; 50.8% of the patients were female; Serum samples were collected at the day of discharge after symptom resolution and SARS-CoV-2 nucleotide testing negative twice by RT-PCR. The median time between the onset of symptom to sample collection was 20 (12-30, IQR) days. The asymptomatic patients were identified by screening those with close-contact history to COVID-19 patients and confirmed by SARS-CoV-2 RT-PCR. The disease severity was stratified into asymptomatic, mild, moderate and severe, according to the national diagnosis and treatment guideline of COVID-19 (7^th^ edition) in China (Supplementary Table 1).

The SARS-CoV-2 spike protein consists of S1 and S2 subunits. The receptor binding domain (RBD) within S1 subunit is essential for virus attachment to host cell receptor, human angiotensin-converting enzyme 2 (ACE2), while S2 is critical for virus entry by mediating viral membrane fusion to host cell membrane^26-28^. Both S1-RBD and S2 represent important potential targets of Nabs^7^. We first compared the antibodies that specifically binding to S1, RBD and S2 of SARS-CoV-2 in sera of COVID-19 recovered patients with different illness severity by IgG ELISA. Notably, severe and moderate symptomatic patients mounted the most and second robust S1, S1-RBD and S2 specific antibodies, respectively, while mild and asymptomatic patients exhibited significantly lower abundances of S1-, S1-RBD- and S2-specific antibodies (Fig.1a-c), highlighting the disease severity as a key determinant factor of the levels of antibodies specific to SARS-CoV-2 S proteins.

Next, we assessed the abilities of antibodies in these COVID-19 recovered patients to block the interaction between RBD and ACE2 Similar to SARS-CoV-2 RBD binding antibodies, the sera from recovered patients with severe symptoms displayed the highest scores of blocking RBD-ACE2 engagement, followed by patients with moderate symptoms, while antibodies from mild and asymptomatic patients showed much inferior capacity to inhibit RBD-ACE2 interaction (Fig. 2a, b).

### Neutralizing antibody responses to SARS-CoV-2 in COVID-19 recovered patients

Subsequently, we conducted SARS-CoV-2 S-protein pseudotyped-lentiviral based neutralization assay to examine the neutralization capacity of sera from COVID-19 recovered patients. Such assay has been proven to be free of biosafety issue but as reliable as the canonical plaque assay with authentic SARS-CoV-2^17,19,21^. We observed that the sera neutralization capacity was positively correlated to disease severity (Fig.2c). Specifically, 80% of patients with severe symptoms and 47.1% of patients with moderate symptoms generated antibodies capable of completely neutralizing pseudovirus infection, while only 25% of serum samples from patients with mild symptoms were able to block pseudovirus infection; strikingly, sera from asymptomatic patients showed no activity to neutralize pseudovirus infection in this assay (Fig. 2d). Next, we performed neutralization assay with the authentic SARS-CoV-2 to explore the neutralization capacity of sera from COVID-19 recovered patients. In accordance with the pseudotyped-lentiviral based neutralization assay, we found that the sera neutralization capacity of severe patients was the highest and was 61.1-fold, 1319.1-fold and 2972.0-fold higher than those of moderate, mild and asymptomatic patients, respectively (Fig. 2e, f).

**Figure 1.**
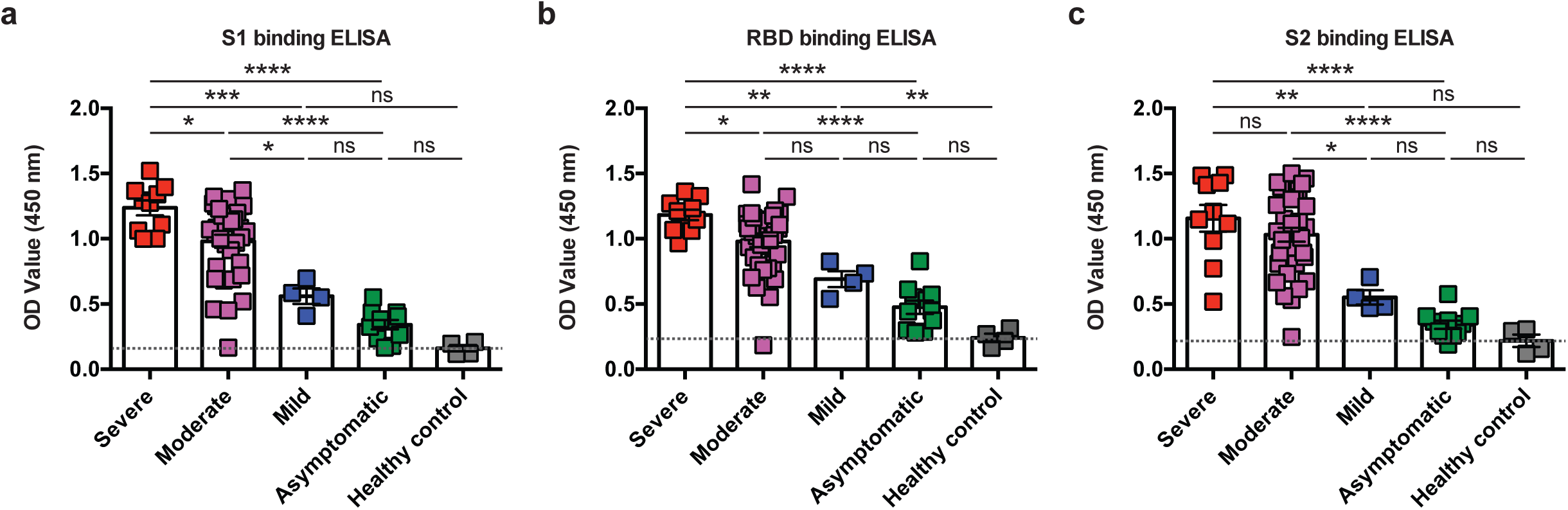
Antibody responses to SARS-CoV-2 in COVID-19 recovered patients with different symptom severity. **a-c**, ELISA binding assays of 100-fold diluted COVID-19 patient sera to ELISA plates coating of SARS-CoV-2 S1 (a), RBD (b) and S2 (c) proteins. The dashed lines in a-c represent the average values of healthy control groups. \**P* < 0.05, \*\**P* < 0.01, \*\*\**P* < 0.001 and \*\*\*\**P* < 0.0001. Not significant, ns. Error bars in a-c indicate SEM.

**Figure 2.**
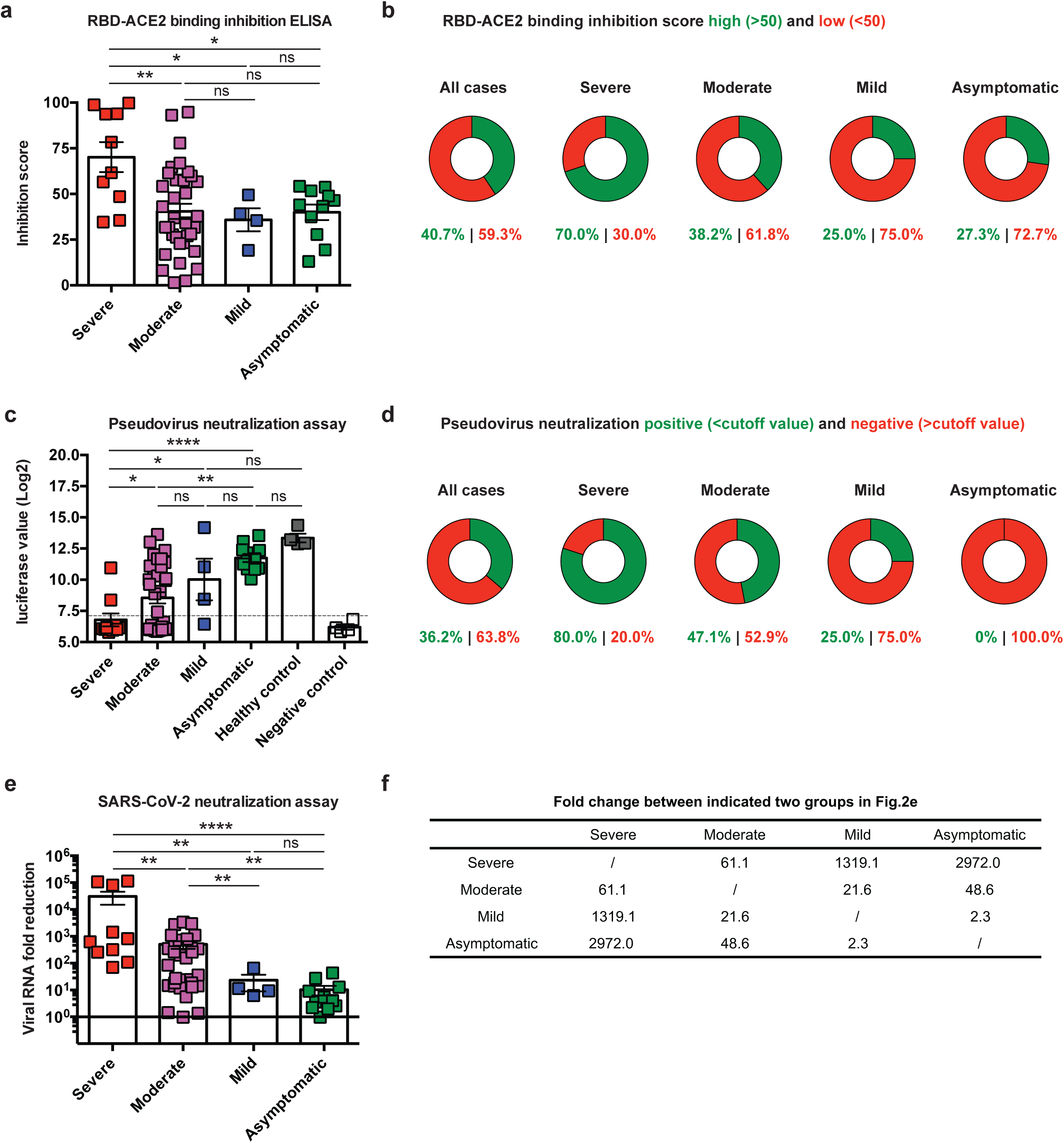
Neutralizing antibody responses to SARS-CoV-2 in COVID-19 recovered patients. **a**, Scores showing the COVID-19 patient serum-mediated inhibition of the SARS-CoV-2 RBD protein binding to ACE2 protein by ELISA. **b**, Pie charts showing the proportions of patients with high (> 50, green) or low (< 50, red) RBD-ACE2 binding inhibition score in each indicated situations. **c**, Patient serum-mediated blocking of luciferase-encoding SARS-CoV-2 typed pseudovirus into ACE2/293T cells. The dashed line indicates the cutoff value (7.121) determined by the ROC curve analysis. **d**, Pie charts showing the proportions of patients with pseudovirus neutralization-positive (< 7.121, green) or -negative (> 7.121, red) in each indicated situations. **e**, Patient serum-mediated blocking of SARS-CoV-2 virus into Vero E6 cells. **f**, A table showing the fold change of SARS-CoV-2 viral RNA fold reduction between indicated two groups in (e). \**P* < 0.05, \*\**P* < 0.01 and \*\*\*\**P* < 0.0001. Not significant, ns. Error bars in a, c, e indicate SEM.

Furthermore, the binding abilities of S1, RBD and S2 were positively correlated to each other (Supplementary Fig. 1a-c) and also positively correlated to both pseudovirus neutralizing capacities (Supplementary Fig. 1d-f) and authentic SARS-CoV-2 virus neutralizing capacities (Supplementary Fig. 1g-i). These data together suggested that the disease severity determines both the magnitude and neutralizing capacity of SARS-CoV-2-specific antibodies in recovered COVID-19 patients.

### Subtypes of neutralizing antibodies to SARS-CoV-2 S proteins in COVID-19 recovered patients

Given that NAbs can potentially target both S1 and S2 to block viral infection^7^, we set out to distinguish S1 and S2-specific NAbs in COVID-19 patients with aforementioned pseudovirus neutralization assay. To this end, we used biotin-labeled S1 or S2 recombinant protein to deplete corresponding antibodies in sera from 25 COVID-19 patients that were confirmed to be highly neutralizing in Fig. 2c and Fig. 2e (Supplementary Fig. 2a, b). After depletion, we found that across all neutralizing sera, 40% of patients generated both competent S1- and S2-NAbs (i.e., post either S1- or S2-specific antibody depletion, sera can still completely neutralize pseudoviruses; labeled as “S1/S2-NAbs”); while 40% of patients only generated S1-competent NAbs (“S1-NAbs only”), and 4% of patients only generated S2-specific NAbs (“S2-NAbs only”); interestingly, 16% of serum samples strictly depended on the collaboration of S1- and S2-specific NAbs to effectively neutralize pseudovirus infection (i.e., either S1- or S2-specific antibody depletion in the serum can result in the failure of neutralization; labeled as “(S1+S2)-NAbs”) (Fig. 3a,b). Among NAbs in severe symptomatic patients, the majority of sera (62.5%) potently neutralized both S1-mediated receptor attachment and S2-mediated membrane fusion, while 37.5% only blocked S1-mediated receptor engagement (Fig. 3c). For mild to moderate symptomatic patients, NAb features were more diverse: 41.2% of them consisted of only S1-neutralizing NAbs, 29.4% possessed the abilities to block both receptor engagement and membrane fusion. Notably, 23.5% of these sera required the combination of S1- and S2-specific NAbs to effectively neutralize pseudovirus infection (Fig. 3c). Collectively, our data revealed the highly heterogeneous nature of NAb responses against SARS-CoV-2 S protein and such diversity seemed to be closely associated with disease severity. The immune mechanisms underlying the diversity of NAbs responses in COVID-19 patients with different degree of symptoms warrant further investigations.

**Figure 3.**
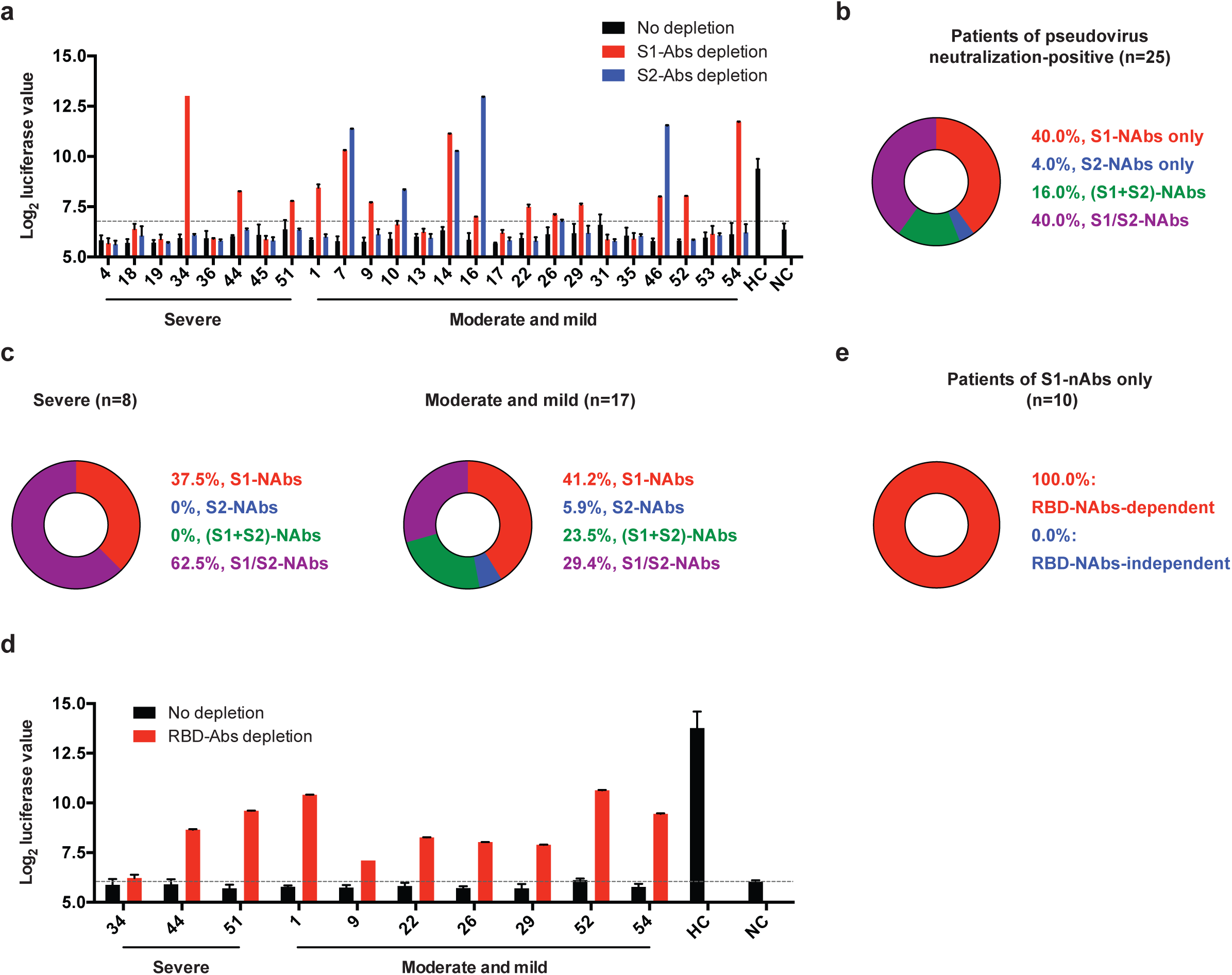
Subtypes of neutralizing antibodies to SARS-CoV-2 S proteins in COVID-19 recovered patients. **a**, Blocking of luciferase-encoding SARS-CoV-2 typed pseudovirus into ACE2/293T cells by patient sera (no depletion) or S1 antibodies-depleted sera (S1-Abs depletion) or S2 antibodies-depleted sera (S2-Abs depletion). The dashed line indicates the cutoff value (6.749) determined by the ROC curve analysis. HC, healthy control. NC, negative control. **b-c**, Pie charts showing the proportions of patients with different neutralizing antibody (NAb) subtype responses in the total 25 patients (b), 8 severe patients (c, left panel) and 17 moderate and mild patients (c, right panel) of pseudovirus neutralization-positive. **d**, Blocking of luciferase-encoding SARS-CoV-2 typed pseudovirus into ACE2/293T cells by “S1-NAbs only” patient sera with RBD antibodies depletion (RBD-Abs depletion) or without RBD antibodies depletion (no depletion). The dashed line indicates the cutoff value (6.034) determined by the ROC curve analysis. HC, healthy control. NC, negative control. **e**, Pie chart showing the proportions of “S1-NAbs only” patients with RBD-NAbs-dependent or -independent antibody response. Error bars in a, d indicate SEM.

Finally, we investigated whether NAbs depleted by S1-recombinant protein are actually targeting RBD for their neutralizing capacity. For this purpose, we depleted RBD-specific antibodies in 10 serum samples showing S1-specific neutralization by biotin-conjugated RBD protein mediated pull-down (Supplementary Fig. 2c). Antibodies post RBD-depletion were shown to lose RBD-binding ability, but still keep their binding to both S1 and S2 proteins, suggesting the efficiency and specificity of RBD Ab depletion (Supplementary Fig. 2c-e). Notably, all sera with S1-specific neutralization failed to neutralize pseudovirus infection after RBD-specific NAb depletion (Fig. 3d, e), demonstrating the strict dependency of RBD-specific NAbs to disengage viral attachment to the host receptor. These data provided the rationale for exclusively using RBD as S1-immunogen in vaccine design, in particular, given that several reports have shown the enhanced disease after whole S1 immunization^9,30^.

## Discussion

The COVID-19 patients show stratified symptoms, including asymptomatic, mild, moderate and severe^2^ Using RBD-ACE2 blockade, pseudovirus neutralization and authentic virus neutralization, we observed that disease severity positively correlates to NAb responses. The patients recovered from severe illness mounted the most robust NAb responses. Strikingly, asymptomatic patients fail to generate competent NAbs. The mechanisms underlying disease severity associated NAb responses are elusive. One possible explanation is that the induction of SARS-CoV-2-specific NAb responses requires the strengthened and prolonged B-cell receptor (BCR) stimulation. Indeed, enhanced BCR rearrangement was observed in COVID-19 patients with severe disease symptom^31^. This may provide important insights into the COVID-19 vaccine design, in which the vaccine regimens should release enough SARS-CoV-2 immunogens in an extended period.

Given the critical role of NAbs in protecting viral infection in airways, the recovered asymptomatic patients may suffer from SARS-CoV-2 re-infection. In this circumstance, these patients need to be vaccinated whenever the effective vaccines are available. Thus far, it is unknown as to the protective immunity that prevents asymptomatic patients from progressing to more severe disease. Probably, these patients can mount robust SARS-CoV-2-specific CD8^+^ T cell responses, which may confer the protection by directly clearing virus-infected target cells. However, this hypothesis needs to be confirmed in future investigations.

Our results also demonstrated the tremendous heterogeneous NAb responses in patients capable of inducing high-titer NAbs. The majority (80.7%) of patients can produce S1-specfic NAbs, and half patients are able to generate S2-specific NAbs. However, only around 40% of patients generated both S1- and S2-specific competent NAbs. Particularly, approximate 7% patients had to depend on the collaboration between S1- and S2-specific antibodies for efficient viral neutralization. The mechanisms underlying the heterogeneous NAb responses in recovered patients remains unknown and warrant further studies. Notably, all S1-specific NAbs were strictly RBD dependent and deletion of RBD-specific antibodies led to the failure in neutralization in S1-specific sera. These results highlighted the importance of S1-RBD itself, but not other parts of S1 protein, in inducing competent NAbs.

In conclusion, we have demonstrated the positive correlation between the magnitude of NAb responses and disease severity in patients recovered from COVID-19. We have also found that disease severity also influences the neutralization heterogeneity of SARS-CoV2-specific antibodies. Our results highlight the needs to include mild-illness and asymptomatic patients for future vaccination, and also suggest the collection of plasma from COVID-19 recovered patients should be restricted to those with moderate to severe symptoms for passive antibody therapy. Our data also provide important rationale for exclusively using SARS-CoV-2 RBD as S1-immunogen in COVID-19 vaccine regime.

## Materials and methods

### Human samples

The 59 COVID-19 recovered patients enrolled in the study were provided written informed consent and from different sources. The sera of the severe, moderate and mild patients were obtained from Guangzhou Eighth People’s Hospital. The sera of the asymptomatic patients were obtained from Chongqing Public Health Medical Center. Healthy control subjects were 4 adult participants in the study. The study received IRB approvals at Guangzhou Eighth People’s Hospital (KE202001134) and Chongqing Public Health Medical Center (2020-023-01-KY).

### ELISA

As previously described^15^, 50 ng of SARS-CoV-2 S1 protein (Sino Biological, 40591-V08H) or SARS-CoV-2 RBD protein (Sino Biological, 40592-V08B) or SARS-CoV-2 S2 protein (Sino Biological, 40590-V08B) in 100 µl PBS per well was coated on ELISA plates (Costar, 42592) overnight at 4°C. The ELISA plates were blocked for 1 hour with 100 µl blocking buffer (5% FBS and 0.1% Tween 20 in PBS) and then incubated with diluted patient or healthy control sera in 100 µl blocking buffer for 1 hour.

After washing with PBST buffer (0.1% Tween 20 in PBS), the ELISA plates were incubated with anti-human IgG HRP antibody (Bioss Biotech, 0297D) for 45 min, followed by PBST washing and addition of TMB buffer (Beyotime, P0209). The ELISA plates were allowed to react for 5-10 min and stopped by 1 M HCl stop buffer. The optical density (OD) value was detected at 450 nm.

### ELISA-based RBD-ACE2 binding inhibition assay

As previously described^15^, 200 ng of ACE2 protein (Sino Biological, 10108-H08H) in 100 µl PBS per well was coated on ELISA plates overnight at 4°C. The ELISA plates were blocked for 1 hour with 100 µl blocking buffer (5% FBS and 0.1% Tween 20 in PBS); meanwhile, 50 µl 10-fold diluted patient or healthy control sera were incubated with 7.5 ng SARS-CoV-2 RBD-mouse FC protein (Sino Biological, 40592-V05H) in 50 µl blocking buffer for 1 hour. Then, the incubated sera/SARS-CoV-2 RBD-mouse FC protein mixture was added into the ELISA plates and allowed to develop for 30 min, followed by PBST washing and incubation with anti-mouse FC HRP antibody (Thermo Fisher Scientific, A16084) for 30 min. Next, the ELISA plates were washed with PBST and treated with TMB buffer (Beyotime, P0209). After 5 min, the ELISA reaction was stopped by 1 M HCl stop buffer and determined at 450 nm. The RBD-ACE2 binding inhibition score was calculated as: 100 × (1 - (OD450 value of patient sera / OD450 value of healthy control sera)).

### Pseudovirus neutralization assay

The pseudovirus neutralization assay was previously described^15,32^.Briefly, HEK-293T cells were transfected with pLenti-luciferase, psPAX2, and 2019-nCov S plasmids by using *Trans*IT-293 Transfection reagent (Mirus, MIR 2700). After 12 hours, the culture media was changed to fresh media. And at 64 hours after transfection, the culture supernatants containing SARS-CoV-2 typed pseudovirus were harvested. Next, 200-fold diluted patient or healthy control sera were mixed with SARS-CoV-2 typed pseudovirus for 1 hour at 37°C. Then, the ACE2-expressing HEK-293T (ACE2/293T) cells were incubated with the sera/pseudovirus mixture overnight and then cultured with fresh media. At 40 hours after the mixture incubation, the luciferase activity of SARS-CoV-2 typed pseudovirus-infected ACE2/293T cells were measured by a luciferase reporter assay kit (Promega, E1910).

### SARS-CoV-2 serum neutralization assay

Patient sera were diluted in DMEM (40 fold-dilution) and mixed with an equal volume of 80-100 PFU SARS-CoV-2 (EPI_ISL_444969) for 1 h at 37°C. Serum-virus mixture were then added to the Vero E6 cell monolayers in 48-well plates and incubated at 37°C in 5% CO_2_ for 1 h. After removing the inocula, plates were overlaid with culture medium and cultured at 37°C for 48 h. Subsequently, viral RNA from the cultural supernatants was extracted and the viral RNA copies were determined by quantitative PCR according to the viral detection kit’s protocol (DAAN Gene Co., Ltd. of Sun Yat-Sen University). All experiments related to authentic viruses were performed in the certified BSL-3 facility of Sun Yat-sen University. The SARS-CoV-2 viral RNA fold reduction = 2^(CT value of sample - CT value of mock)^.

### Depletion of SARS-CoV-2 S protein-specific antibodies

Firstly, SARS-CoV-2 S1 protein (Sino Biological, 40591-V08H) or SARS-CoV-2 RBD protein (Sino Biological, 40592-V08B) or SARS-CoV-2 S2 protein was conjugated with biotin by following the manufacture’s protocol (Thermo Fisher Scientific, A39257). Then, biotin-conjugated proteins were incubated with BeaverBeads Mag Streptavidin Matrix (Beaver, 22305) at 4°C for 1.5 hours. After washing with PBS, the SARS-CoV-2 S protein coupled beads were next incubated with diluted patient sera at 4°C for 1.5 hours. Then, the supernatants were harvested and quality controlled by ELISA assays for SARS-CoV-2 S proteins.

### Statistics

The SARS-CoV-2 antibody titers or virus neutralizing function of the sera belonging to patients with different severity were compared with the one-way ANOVA test (Tukey’s multiple comparisons test). The cutoff value in each pseudovirus neutralizing function assay was determined by the ROC curve analysis and was of the highest likelihood ratio. Correlations between different SARS-CoV-2 antibody titers or between SARS-CoV-2 antibody titers and pseudovirus titers or between SARS-CoV-2 antibody titers and SARS-CoV-2 virus titers were analyzed using Pearson’s correlation coefficient. *P* values less than 0.05 were defied was statistically significant. Prism 6 software was used for statistical analysis.

## Data Availability

There are no additional data in the study.

## Data availability statements

The data sets in the study are available from the corresponding authors upon reasonable request.

## Acknowledgements

This work was supported by grants from the National Science and Technology Major Project (No. 2017ZX10202102-006-002 to L.Y.), the National Natural Science Fund for Distinguished Young Scholars (No. 31825011 to L.Y.) and the Chongqing Special Research Project for Novel Coronavirus Pneumonia Prevention and Control (No. cstc2020jscx-2 to L.Y.; No. cstc2020jscx-fyzx0074 to Y.C.; cstc2020jscx-fyzx0135 to Y.C.).

## Contributions

X.C., Z.P., S.Y., F.Y., J.Z., Y.Y., R.L., B.L., X.Y., L.G., Z.L., Y.L., Q.H., L.X., J.T., L.H. and J.Z. performed the experiments. L.Y. designed the study, analyzed the data and wrote the paper with X.C., X.Z., P.L., Y.W. and K.D.; and G.Z., Y.C., K.D. and L.Y. supervised the study.

---

**Supplementary Figure 1.**
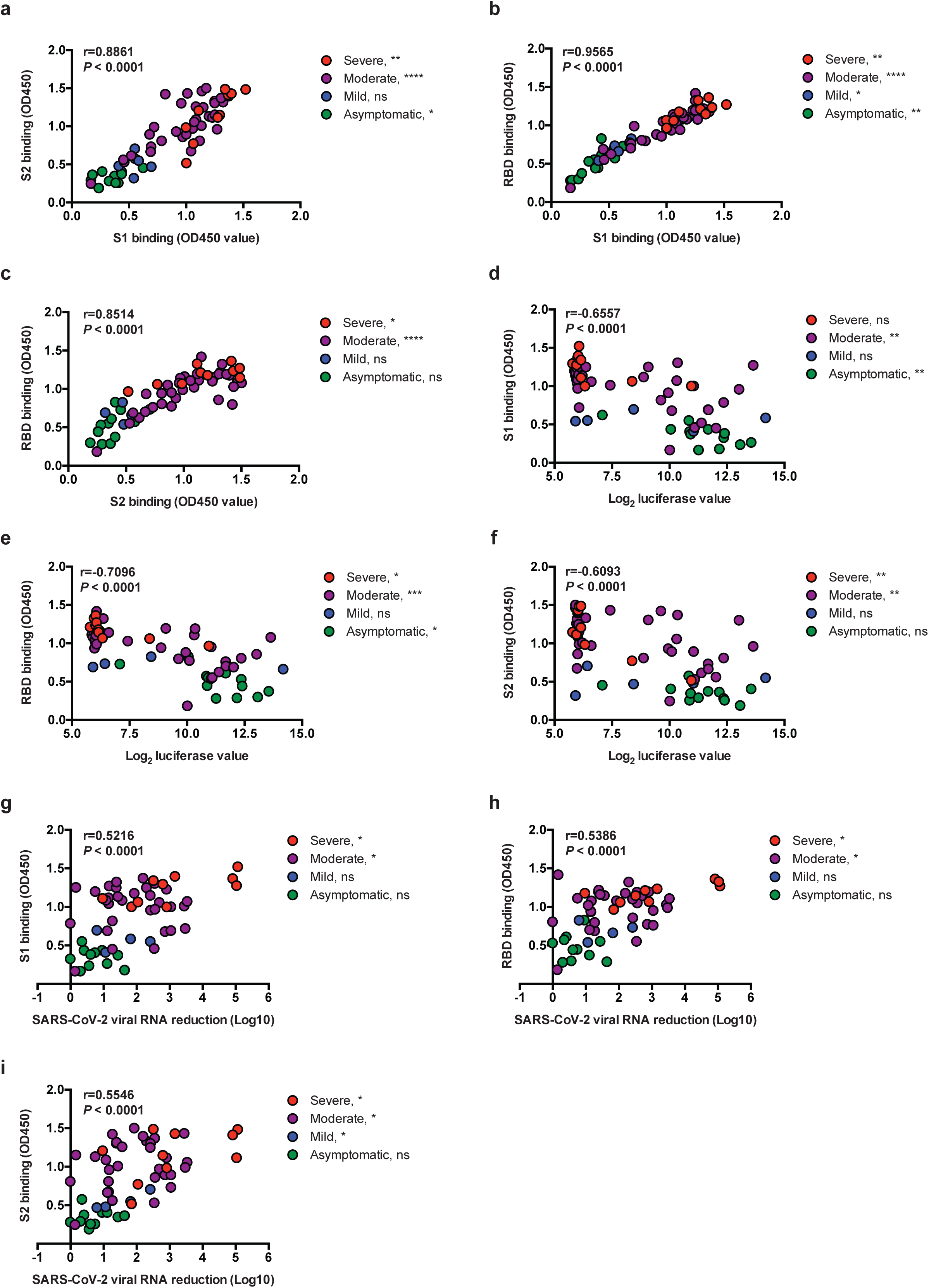
Correlation analyses of SARS-CoV-2 antibody titers and neutralizing functions in COVID-19 recovered patients. **a-c**, Correlations between SARS-CoV-2 S1 antibody titer and SARS-CoV-2 S2 antibody titer (a), SARS-CoV-2 S1 antibody titer and SARS-CoV-2 RBD antibody titer (b) and SARS-CoV-2 S2 antibody titer and SARS-CoV-2 RBD antibody titer (c) in the 59 COVID-19 recovered patients. **d-f**, Correlations between SARS-CoV-2 S1 antibody titer and SARS-CoV-2 pseudovirus neutralizing function (d), SARS-CoV-2 RBD antibody titer and SARS-CoV-2 pseudovirus neutralizing function (e) and SARS-CoV-2 S2 antibody titer and SARS-CoV-2 pseudovirus neutralizing function (f) in the 59 COVID-19 recovered patients. **g-i**, Correlations between SARS-CoV-2 S1 antibody titer and SARS-CoV-2 virus neutralizing function (g), SARS-CoV-2 RBD antibody titer and SARS-CoV-2 virus neutralizing function (h) and SARS-CoV-2 S2 antibody titer and SARS-CoV-2 virus neutralizing function (i) in the 59 COVID-19 recovered patients. The *P* value and r value at the top left of each panel assess the correlation between the indicated two parameters in the total 59 patients. The asterisks adjacent to severity at the top right of each panel assess the correlation between the indicated two parameters in patients of indicated severity. \**P* < 0.05, \*\**P* < 0.01, \*\*\**P* < 0.001 and \*\*\*\**P* < 0.0001. Not significant, ns.

**Supplementary Figure 2.**
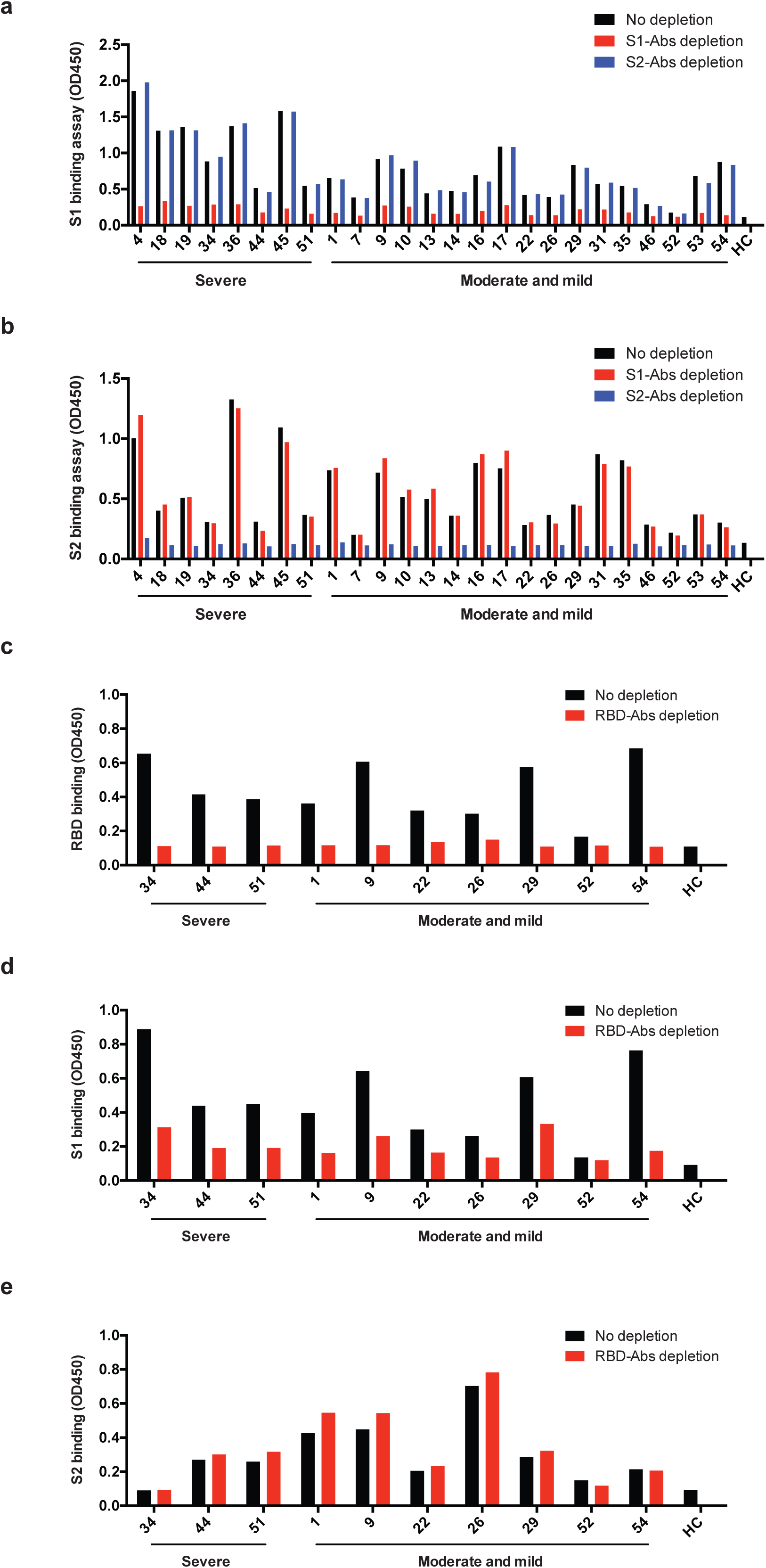
Depletion efficiencies of SARS-CoV-2 S1, S2 and RBD antibodies in patient sera. **a, b**, ELISA binding assays of 1500-fold diluted COVID-19 patient sera (no depletion) or S1 antibodies-depleted sera (S1-Abs depletion) or S2 antibodies-depleted sera (S2-Abs depletion) to ELISA plates coating of SARS-CoV-2 S1 (a) and S2 (b) proteins. HC, healthy control. **c-e**, ELISA binding assays of 1500-fold diluted COVID-19 patient sera (no depletion) or RBD antibodies-depleted sera (RBD-Abs depletion) to ELISA plates coating of SARS-CoV-2 RBD (c), S1 (d) and S2 (e) proteins. HC, healthy control.

**Supplementary Table 1.** Characteristics of patients from the study. 1. Continuous variables were shown as the median (interquartile range, IQR). 2. All the 59 COVID-19 patients were tested positive qPCR for SARS-CoV-2 virus RNA upon hospital admission. Patients were diagnosed as severe when meeting any one of the following conditions: 1) anhelation (RR ≥ 30/min), 2) SpO2 ≤ 93%, 3) PaO2/FiO2 ≤ 300 mmHg and 4) imageological diagnosis of signigicant progress (> 50%) in 24-48 hours. The moderate patients were diagnosed with respiratory symptoms, fever and imageological evidence of pneumonia. The mild patients were diagnosed with mild clinical symptoms and no imageological evidence of pneumonia. The asymptomatic patients were those without clinical symptoms.

|  | All (n=59) | Severe (n=10) | Moderate (n=34) | Mild (n=4) | Asymptomatic (n=11) |
| --- | --- | --- | --- | --- | --- |
| Age-years (median, IQR) | 47 (33-62) | 64 (58-70) | 46 (34-62) | 21 (20-32) | 42 (24-49) |
| Gender |  |  |  |  |  |
| Female | 30 (50.8%) | 4 (40.0%) | 18 (52.9%) | 4 (100.0%) | 4 (36.4%) |
| Male | 29 (49.2%) | 6 (60.0%) | 16 (47.1%) | 0 (0%) | 7 (63.6%) |
| Onset of symptom to sample collection-days (median, IQR) | 20 (12-30) | 17 (14-21) | 20 (10-29) | 24 (20-27) | 23 (20-32) |
| Hospital admission date (medium, IQR) | 2020/1/30<br>(2020/1/25 to 2020/3/13) | 2020/1/26<br>(2020/1/24 to 2020/1/28) | 2020/1/27<br>(2020/1/25 to 2020/2/3) | 2020/3/22<br>(2020/3/11 to 2020/3/23) | 2020/4/9<br>(2020/3/27 to 2020/4/14) |

